# Assessment of private variants in *PRKN, PARK7* and *PINK1* in Parkinson’s disease

**DOI:** 10.1101/2022.02.18.22269402

**Authors:** Jing Hu, Cheryl H. Waters, Dan Spiegelman, Edward A. Fon, Eric Yu, Farnaz Asayesh, Lynne Krohn, Prabhjyot Saini, Roy N. Alcalay, Sharon Hassin-Baer, Ziv Gan-Or, Dimitri Krainc, BaoRong Zhang, Bernabe I. Bustos, Steven J. Lubbe, the International Parkinson’s Disease Genomics Consortium (IPDGC)

## Abstract

Recessive mutations in *PRKN, PARK7* and *PINK1* are established causes of early-onset Parkinson’s disease (EOPD). Previous studies have interrogated the role of heterozygous variants in these genes but mainly focused on rare (minor allele frequency [MAF] <1%) damaging variants or established mutations. Here, we assessed heterozygous private *PRKN, PARK7* and *PINK1* variants in PD risk in four large-scale PD case-control datasets by performing gene-wise burden analyses using sequencing data totaling 5,829 PD cases and 7,221 controls, and summary allele counts from 9,501 PD cases and 48,207 controls. Results showed no significant burden in all three genes after meta-analyses. Burden in EOPD (age at onset <50 years) and late-onset PD (≥50 years) remained nonsignificant. In summary, we found no evidence to support the association of the excess burden of heterozygous private variants in *PRKN, PARK7*, and *PINK1* with PD risk in the European population. Larger, more diverse cohorts are needed to accurately determine their role in PD.

## Introduction

Biallelic variants in *PRKN* (*Parkin*), *PARK7* (*DJ-1*) and *PINK1* cause early-onset Parkinson’s disease (EOPD, defined here as age at onset [AAO] <50 years) (Bonifati, 2012; Kasten et al., 2018; Kilarski et al., 2012). The role of heterozygous mutations is more controversial. Several recent large-scale studies did not uncover any association for heterozygous mutations in these genes with PD (Krohn et al., 2020; Lubbe et al., 2021; Yu et al., 2021; Zhu et al., 2021). These studies focused on rare (minor allele frequency [MAF] <1%) damaging variants or established mutations, and largely ignored the specific role of private variants that are only seen in single individuals within a cohort and are absent or very rare in public population databases such as gnomAD (https://gnomad.broadinstitute.org; (Karczewski et al., 2020)). An increased burden of rare (MAF <1%) and private heterozygous *PRKN* variants in late-onset PD (LOPD, AAO ≥50 years) was recently reported (Hopfner et al., 2020). Here, we analyzed heterozygous private variants in *PRKN, PARK7* and *PINK1* using four large PD cohorts to assess their role in PD risk.

## Methods

Sequencing data were obtained from three independent PD case-control cohorts totaling 5,829 PD cases and 7,221 controls, and summary count data from 9,501 PD cases and 48,207 controls was extracted from the *PD Variant Browser* (Kim et al., 2021) and two of the sequencing cohorts (International Parkinson’s disease Genomics Consortium (IPDGC)-whole exome sequencing (WES) and McGill). Cohort information is described in **Supplementary Table 1**. Heterozygous private *PRKN, PARK7* and *PINK1* variants were prioritized using, as a guideline, a MAF<1% threshold in >141,000 population-based individuals (Karczewski et al., 2020). Damaging private variants were identified as described in **Supplementary Methods**. To avoid overlap between the sequencing cohorts and those used to extract allele counts, different analytical methods were used. R package *MetaSKAT* (Lee et al., 2013) and *Metafor* (Viechtbauer, 2010) were utilized for meta-analysis of sequencing data and allele counts data, respectively. Bonferroni correction based on the number of genes was applied for multiple testing. See **Supplementary Methods** for more details.

## Results

Using available sequencing data, we first performed a gene-based Optimized Sequence-kernel-association test (SKAT-O) (Lee et al., 2012) to assess the burden of all heterozygous private and private damaging variants that yielded no significant association for all genes in individual cohort analysis, nor after meta-analysis (**Table 1**). Similarly, after stratifying PD cases into EOPD and LOPD, no significant enrichment was observed (**Supplementary Tables 2 and 3**).

**Table 1.**
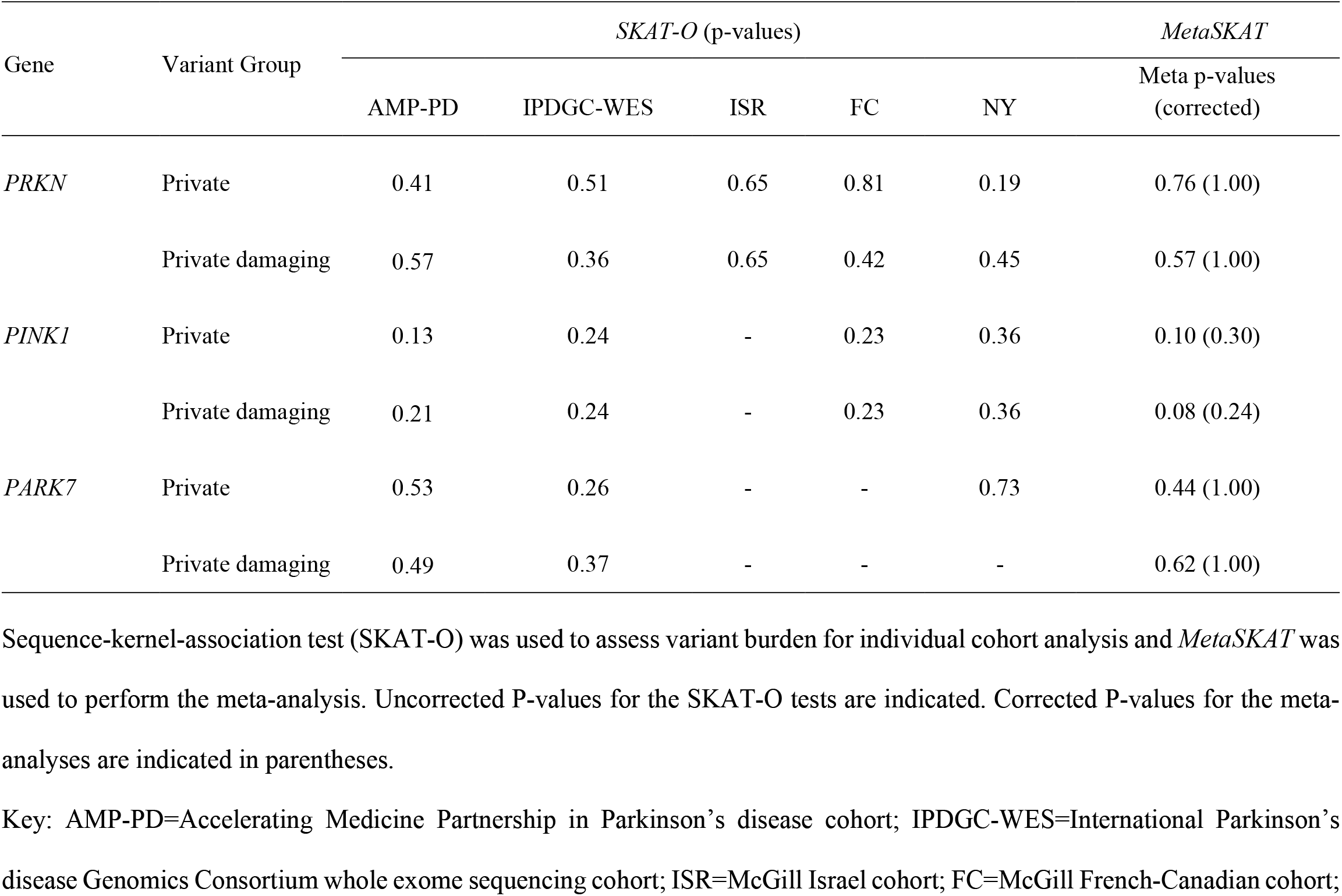
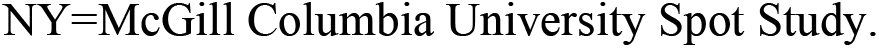
Assessment of private variants in *PRKN, PARK7* and *PINK1* with Parkinson’s disease (PD) in cohorts with available sequencing data.

| Gene | Variant Group | *SKAT-O* (p-values) |  |  |  |  | *MetaSKAT* |
| --- | --- | --- | --- | --- | --- | --- | --- |
|  |  | AMP-PD | IPDGC-WES | ISR | FC | NY | Meta p-values (corrected) |
| *PRKN* | Private | 0.41 | 0.51 | 0.65 | 0.81 | 0.19 | 0.76 (1.00) |
|  | Private damaging | 0.57 | 0.36 | 0.65 | 0.42 | 0.45 | 0.57 (1.00) |
| *PINK1* | Private | 0.13 | 0.24 | - | 0.23 | 0.36 | 0.10 (0.30) |
|  | Private damaging | 0.21 | 0.24 | - | 0.23 | 0.36 | 0.08 (0.24) |
| *PARK7* | Private | 0.53 | 0.26 | - | - | 0.73 | 0.44 (1.00) |
|  | Private damaging | 0.49 | 0.37 | - | - | - | 0.62 (1.00) |

Sequence-kernel-association test (SKAT-O) was used to assess variant burden for individual cohort analysis and *MetaSKAT* was used to perform the meta-analysis. Uncorrected P-values for the SKAT-O tests are indicated. Corrected P-values for the meta-analyses are indicated in parentheses.
Key: AMP-PD=Accelerating Medicine Partnership in Parkinson’s disease cohort; IPDGC-WES=International Parkinson’s disease Genomics Consortium whole exome sequencing cohort; ISR=McGill Israel cohort; FC=McGill French-Canadian cohort;
NY=McGill Columbia University Spot Study.

To further assess the contribution of *PRKN, PARK7* and *PINK1* heterozygous private variants, we next extracted allele counts and performed an allelic gene-based Fisher’s exact burden test. Again, no significant associations at the individual cohort level were observed for both private and private damaging variants (**Supplementary Table 4**). After meta-analysis, no significant results were found for private variants (**Supplementary Figure 1**) and private potentially damaging variants (**Figure 1**) in the three genes. After stratifying PD cases into EOPD and LOPD, private and private potentially damaging *PRKN* variants demonstrated some level of enrichment in the McGill EOPD New York (NY) cohort (**Supplementary Table 4**), however, these findings no longer remained after meta-analysis (**Supplementary Figures 2-5)**.

**Figure 1.**
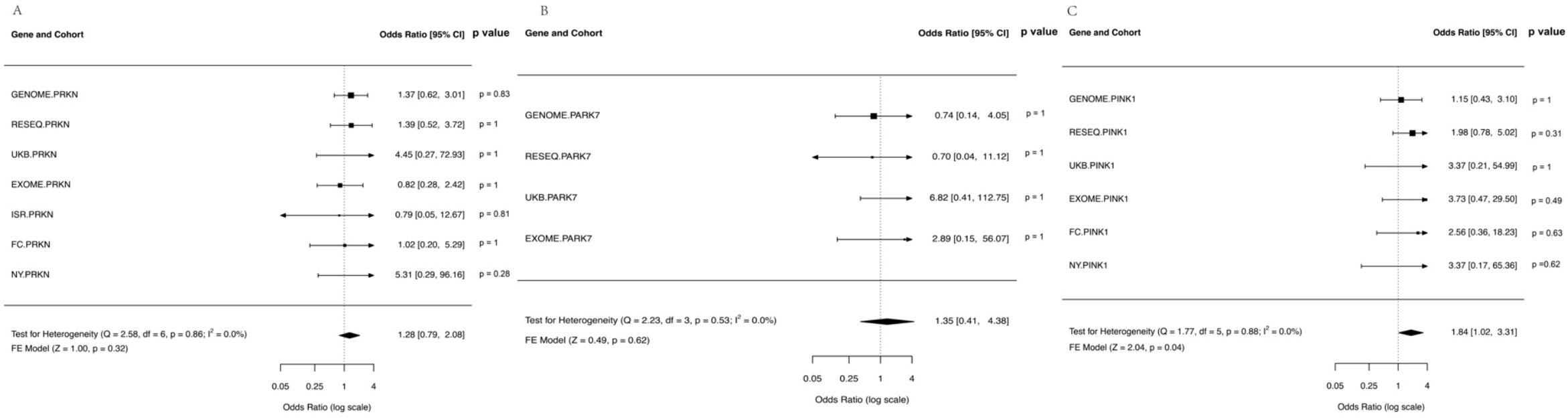
Meta-analysis of carriership burden of private damaging variants in *PRKN, PARK7* and *PINK1* genes in Parkinson’s disease (PD) using allele count data. Forest plots assessing private, potentially damaging variants (CADD >20) in (A) *PRKN* (*Parkin*), (B) *PARK7* (*DJ-1*) and (C) *PINK1* genes. Allele counts for each gene were extracted from the *PD Variant Browser*, International Parkinson’s disease Genomics Consortium (IPDGC) Whole Exome Sequencing Project cohort and individual McGill cohorts separately. Meta-analyses were performed using *Metafor*. Panels show Odds ratio, confidence intervals (CI), corrected Fisher’s exact test gene burden p-value per cohort (right side of each plot), and the overall meta-analysis p-value per gene (bottom left of each plot). Key: CADD=*Combined Annotation Dependent Depletion* phred-scaled score; GENOME=PD Genome Project cohort; RESEQ=IPDGC Resequencing Project cohort; UKB=UK Biobank cohort; EXOME=IPDGC Whole Exome Sequencing Project cohort; ISR=McGill Israel cohort; FC=McGill French/French-Canadian cohort; NY=McGill Columbia University Spot Study; FE=fixed effects; I^2^=I-square test for heterogeneity.

## Discussion

Here we assessed the contribution of heterozygous private variants within *PRKN, PARK7* and *PINK1* to PD risk in several large European ancestry cohorts. Overall, no enrichment across all genes was observed for individual cohorts and upon meta-analysis for both the sequencing and allele count data, and also not after stratifying by early and late onset PD cases. Together this suggests that private *PRKN, PARK7* and *PINK1* variants do not influence PD risk in the European population. While in agreement with previous studies assessing heterozygous mutations (Krohn et al., 2020; Lubbe et al., 2021; Yu et al., 2021; Zhu et al., 2021), our findings are not consistent with those of Hopfner et al. (Hopfner et al., 2020). Differences in genotyping methodologies (pooled DNA sequencing and individual genotype simulation compared to whole-exome, genome, or targeted sequencing in our case) could explain this discordance (see **Supplementary Discussion**). Additionally, cohort characteristics and failure to assess all possible private variants, which can influence *PRKN* burden assessments (Krohn et al., 2020; Lubbe et al., 2021; Yu et al., 2021; Zhu et al., 2021), may also play a role. Despite our study being well powered to detect significant gene-based variant enrichment (power=85.7%; α=0.05; based on 3.4-fold enrichment and gene-based carriership of 0.03% in controls (Hopfner et al., 2020) and global disease prevalence of 0.2% (Keller et al., 2012)), we failed to uncover evidence that private *PRKN, PARK7* and *PINK1* variants influence PD risk in the European population. Additional studies of private variants in more diverse populations are needed to further understand their role in PD.

## Data Availability

All genetic data used in this study is available (upon application) from the following sites:
(i) Accelerating Medicines Partnership: Parkinson's Disease (https://amp-pd.org).
(ii) International Parkinson's Disease Genomics Consortium (https://pdgenetics.org/resources).
(iii) Parkinson's Disease Variant Browser (https://pdgenetics.shinyapps.io/VariantBrowser).
(iv) Dataset using targeted next-generation sequencing with Molecular inversion probes (https://cbigr-open.loris.ca/)

