## Supplementary for "Assessment of private variants in *PRKN, PARK7* and *PINK1* in Parkinson’s disease"

### Supplementary Methods

#### Subjects

*(i) The Accelerating Medicine Partnership in Parkinson's disease (AMP-PD) whole genome sequencing (WGS) cohort.*

The AMP-PD cohort includes Michael J. Fox Foundation (MJFF) and National Institutes of

Neurological Disorders and Stroke (NINDS) BioFIND study, Harvard Biomarkers Study (HBS), the NINDS Parkinson's disease Biomarkers Program (PDBP), MJFF Parkinson's Progression Marker Initiative (PPMI), NIA International Lewy Body Dementia Genetics Consortium Genome Sequencing in Lewy body dementia case-control cohort (LBD), the MJFF *LRRK2* Cohort Consortium (LCC) cohort, and the NINDS Study of Isradipine as a Disease Modifying Agent in Subjects With Early Parkinson Disease, Phase 3 (STEADY-PD3). It is composed of 2,494 PD cases and 3,559 healthy controls, corresponding to the release v.2. Among PD cases, 206 (8%) are early-onset Parkinson's disease (EOPD, defined as age at onset [AAO] <50 years) and 2,288 (92%) are late-onset Parkinson's disease (LOPD, defined as age at onset [AAO] ≥50 years). See **Supplementary Table 1**. Generation, processing details and quality controls have been described in the AMP-PD portal (https://amp-pd.org). Further filtering to the VCF files was done, removing variants with read depth (DP) < 20 or genotype quality (GQ) < 20.

*(ii) International Parkinson's Disease Genomics Consortium (IPDGC) whole exome sequencing (WES) cohort (IPDGC-WES).*

The IPDGC WES data consisted of 1,108 PD cases and 456 healthy controls (https://pdgenetics.org/resources). Among PD cases, 584 (53%) are EOPD, 76 (7%) are LOPD, and 448 (40%) cases lack the age information. See **Supplementary Table 1.** The generation, processing details and quality controls are previously described (Jansen et al., 2017).

*(iii) Dataset using targeted next-generation sequencing with Molecular inversion probes (McGill)*

This dataset includes three cohorts, a European ancestry cohort of French-Canadian and French participants recruited at McGill University (FC cohort, 910 PD cases and 2,330 healthy controls), a self-reported European and Ashkenazi Jewish ancestry cohort collected at Columbia University, New York (Columbia University Spot (NY) cohort, 893 PD cases and 428 healthy controls) and a cohort of self-reported Ashkenazi Jewish individuals from Israel (ISR cohort, 661 PD cases and

523 healthy controls). Among PD cases, 160 (18%) are EOPD and 566 (62%) are LOPD in the FC cohort; 92 (10%) are EOPD and 801 (90%) are LOPD in the NY cohort; 125 (19%) are EOPD and 513 (78%) are LOPD in the ISR cohort. See **Supplementary Table 1**. The generation, processing details and quality controls have been described previously (Alcalay et al., 2015; Gan-Or et al., 2020; Kim et al., 2021; Ruskey et al., 2019; Yu et al., 2021).

*(IV) Parkinson's Disease Variant Browser*

Allele counts were extracted from the *PD Variant Browser* (Kim et al., 2021), which is a publicly available database that contains >6 million variants collated from (i) UK Biobank (114 PD cases, 1,650 proxy cases and 38,263 healthy controls), (ii) PD Genome Project (GENOME, 2,744 PD cases and 4,071 healthy controls), (iii) IPDGC Exomes (2,110 PD cases and 2,978 healthy controls), (iv) IPDGC Resequencing project (RESEQ, 3,071 PD cases and 2,136 healthy controls) and (v) IPDGC GWAS Cohort (21,412 PD cases and 23,894 healthy controls) (https://pdgenetics.shinyapps.io/VariantBrowser/). This browser included part of the AMP-PD WGS and IPDGC-WES cohorts mentioned above.

### Variant annotation and selection

Variants were annotated using *ANNOVAR* (Wang et al., 2010) and the *Combined Annotation Dependent Depletion* (CADD) phred-scaled score (https://cadd.gs.washington.edu) (Rentzsch et al., 2021). Private missense, splice-site and stop-gain variants in *PRKN*, *PARK7* and *PINK1* with a minor allele frequency (MAF) <1% in the Genome Aggregation Database (gnomAD v3.1.1, https://gnomad.broadinstitute.org) (Karczewski et al., 2020) were selected. Damaging variants were defined as follows: missense, splice-site and stop-gain variants with a stringent CADD phred-scaled score >20, representing the top 2% predicted deleterious variants.

### Gene wise burden analysis and Fisher’s exact test

Cohort information is described in **Supplementary Table 1**. We leveraged genotypes from three PD case-control sequencing cohorts, including AMP-PD WGS, IPDGC WES and McGill, to assess heterozygous private variant burden in *PRKN*, *PARK7* and *PINK1* using the Optimized Sequence-kernel-association test (SKAT-O) (Lee et al., 2012). Age, sex, 10 principal components, *GBA* status, and ethnicity were used as covariates, if available. Fisher's exact tests were performed on the collapsed variant counts per gene to assess the carriership burden using summary counts data from the *PD Variant Browser*, IPDGC WES, and McGill data. AMP-PD WGS data were excluded from the burden test using summary allelic counts as they are part of the *PD Variant Browser* cohort. Furthermore, the IPDGC Exomes cohort from *PD Variant Browser* was also excluded from the burden test using summary allelic counts due to the high missing rate of the tested variants over the genes of interest, while the array data of IPDGC GWAS Cohort from *PD Variant Browser* was removed as well due to the difficulty of imputing private variants as not all variants can be interrogated given the preselected nature. For the EOPD and LOPD analyses, we utilized 2,917 cases and 3,737 controls in the total allele count data, 5,381 cases and 7,221 controls in the sequencing data with available AAO information. Individual cohort burden analysis and meta-analysis were both performed.

**Meta-Analysis**

The sequencing data from AMP-PD WGS, IPDGC-WES and McGill cohorts were leveraged for SKAT-O meta-analysis using the R package *MetaSKAT* (Lee et al., 2013). Allele counts from the *PD Variant Browser*, IPDGC-WES and McGill cohorts were utilized for meta-analysis with R package *Metafor* (Viechtbauer, 2010). In *Metafor*, heterogeneity was assessed by $I^2$ (total heterogeneity / total variability) = 100% x (Q - df) / Q, where Q denotes the test statistic for heterogeneity and df is the degrees of freedom of the test.

**Supplementary Results**

Cohort information is described in **Supplementary Table 1**. Biallelic mutation carriers in all three genes were removed before burden analyses: one case with a homozygous variant p.Ser65Asn in *PRKN* in GENOME cohort; one case with a homozygous variant p.Met458Leu in *PRKN* in RESEQ cohort; one case with a homozygous variant p.Arg42Pro in *PRKN* in RESEQ cohort; one case with a homozygous variant p.Leu347Pro in *PINK1* in the NY cohort; and one case with compound heterozygotes p.Arg420His + p.Pro37Leu in *PRKN* in the NY cohort.

Meta-analysis of available sequencing data for all three genes yielded no significant association of private or private damaging variants after stratification into EOPD cases (**Supplementary Table 2**) and LOPD cases (**Supplementary Table 3**).

Meta-analysis of allele counts of private variants for all three genes is shown in **Supplementary Figure 1**.

Meta-analysis of allele count data of private and private damaging variants for all three genes in EOPD are shown in **Supplementary Figures 2 and 3** respectively and for LOPD in **Supplementary Figures 4 and 5** respectively.

Overall, meta-analyses of extracted allele count data failed to identify significant enrichment of private variants and private damaging variants for *PARK7* and *PINK1*. A detail including the numbers of carrier in cases, carrier in controls, noncarrier in cases and noncarrier in controls, raw p value and the corrected p value from Fisher's exact tests using allele count data for each separate cohort was described in **Supplementary Table 4**. Both private (OR=23.67, 95% CI: 1.13-497.3, Pcorr=0.03) and private damaging *PRKN* variants (OR=23.67, 95% CI: 1.13-497.3, Pcorr=0.03) were demonstrated some level of enrichment in the EOPD NY cohort after multiple testing correction (**Supplementary Table 4**). Upon inspection, the same variants were present in the private and private damaging variant *PRKN* groups. These associations disappeared upon meta-analysis (**Supplementary Figures 2-3)**. Note that the meta-analysis for the *PARK7* gene failed due to limited cohorts with alternative counts after stratification into EOPD and LOPD cases, but Fisher's exact test of each individual cohort yielded no significance (see **Supplementary Table 4**).

Information regarding the numbers of cases/controls with homozygous A1 alleles, heterozygous alleles, homozygous A2 alleles, amino acid change, *etc*., for each private variant or private damaging variant in individual cohorts from *PD Variant Browser* is detailed in **Supplementary Tables 5 and 6**, respectively. Details of the private and private damaging variants leveraged by burden analyses, including chromosome number, position, reference allele, alternative allele, allele frequency in gnomAD, amino acid change, *etc*., for all the sequencing cohorts are shown in **Supplementary Tables 7 and 8**, respectively.

**Supplementary Discussion**

Our study assessed the contribution of private variants in *PRKN*, *PARK7* and *PINK1* in PD risk in the European population. Overall we did not identify any enrichment of private or private damaging variants within the three genes in several large PD case-control cohorts of European ancestry. However, after AAO stratification, some evidence for enrichment of heterozygous private *PRKN* variants was seen in EOPD in the NY cohort using allele count data. Caution is needed when interpreting this finding due to the small number of EOPD cases in the NY cohort (n=93). This finding was subsequently lost after a meta-analysis of several cohorts totaling 961 EOPD cases and 3,737 controls. Additional large cohorts are needed to further interrogate the role of private and private damaging *PRKN* variants.

Hopfner et al. identified enrichment of rare and private *PRKN* variants in LOPD (Hopfner et al., 2020). Our results do not support this, and the different approaches used in determining variant genotypes could explain this. In order to overcome the lack of individual genotypes required by the SKAT-O, Hopfner et al. simulated individual level genotypes on the basis of observed minor allele counts in each pool from pooled DNA sequencing. False rare variants are the most challenging issue with this approach, possibly generating false positives (Anand et al., 2016), which may lead to potential bias therefore cautious interpretation of their results is needed. Similarly, the identification of rare variants from next generation sequencing (NGS) data can be influenced by various quality controls employed, for example selecting variants with a minimum read depth (DP). Although we used established standards for variant identification from the data available to us, we may have inadvertently missed some private variants - but this would have been consistent across both cases and controls. Our study analyzes some of the largest NGS PD datasets currently available and is well powered to detect significant gene-based private variant enrichment (power=85.7%); however, AAO data was only available for 2,917 cases and 3,737 controls of the summary counts data used which resulted in reduced power to detect meaningful links between private variants and AAO in PD. Additional studies in larger cohorts are needed to elucidate the role (if any) of private variants in influencing AAO of PD.

In conclusion, we failed to uncover any evidence in support of the role of private variants in *PRKN*, *PARK7,* and *PINK1* in PD within European populations. Additional studies of private variants in more diverse populations are needed to further understand their role in PD.

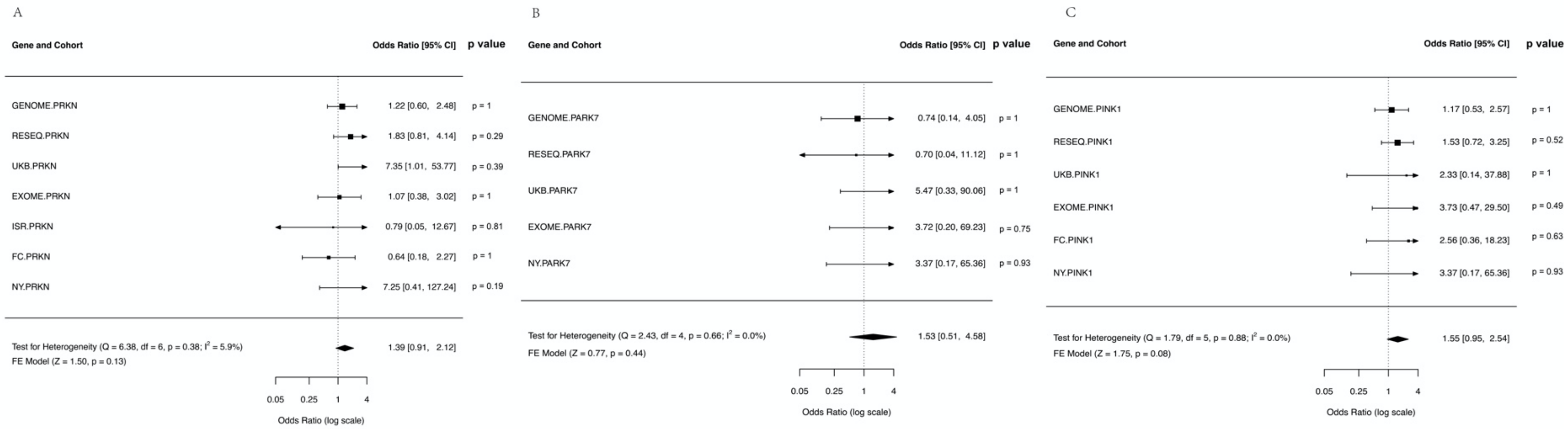


**Supplementary Figure 1. Meta-analysis of carriership burden of private variants in *PRKN, PARK7 and PINK1* genes in PD using allele count data**.

Forest plot of (A) *PRKN* (*Parkin*), (B) *PARK7* (*DJ-1*) and (C) *PINK1* per cohort in rare private variants group. Allele counts for each gene were extracted from the PD Variant Browser, IPDGC Whole Exome Sequencing Project cohort and McGill cohort separately. Meta-analyses were performed using *Metafor*. Panels show Odds ratio, confidence intervals (CI), corrected Fisher's exact test gene burden p-value per cohort (right side of each plot), and the overall meta-analysis p-value per gene (bottom left of each plot).

Key: GENOME=PD Genome Project cohort; RESEQ=IPDGC Resequencing Project cohort; UKB=UK Biobank cohort; EXOME=IPDGC Whole Exome Sequencing Project cohort; ISR=McGill Israel cohort; FC=McGill French-Canadian cohort; NY=McGill Columbia University Spot Study; FE=fixed effects; $I^2$=I-square test for heterogeneity.

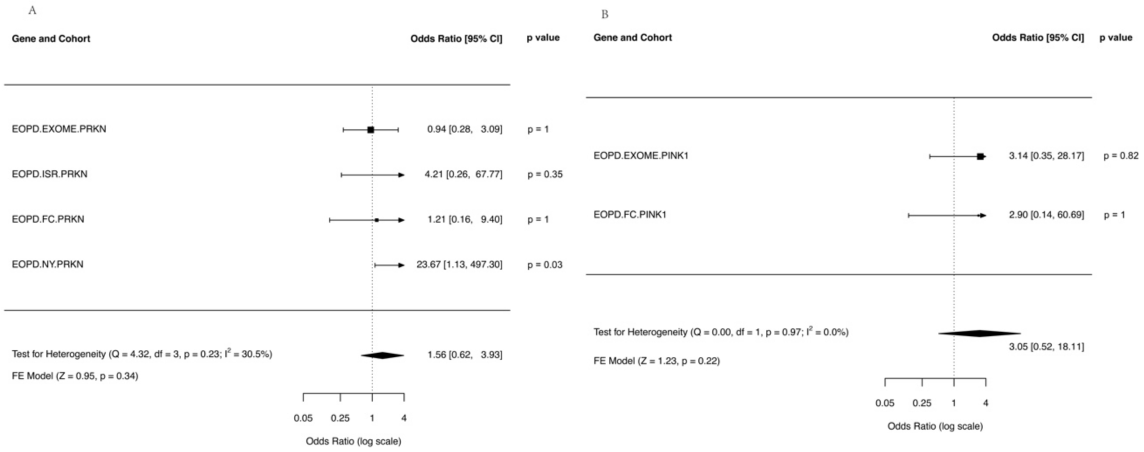


**Supplementary figure 2. Meta-analysis of carriership burden of private variants in *PRKN, and PINK1* genes in the EOPD cohort using allele count data**.

Forest plot of (A) *PRKN* (*Parkin*), (B) *PINK1* per cohort in rare private variants group in the EOPD cohort. Allele counts for each gene were extracted from the IPDGC Whole Exome Sequencing Project cohort and McGill cohort separately. Meta-analyses were performed using *Metafor*. Panels show Odds ratio, confidence intervals (CI), corrected Fisher's exact test gene burden p-value per cohort (right side of each plot), and the overall meta-analysis p-value per gene (bottom left of each plot).

Key: EOPD=Early-onset Parkinson's disease; EXOME=IPDGC Whole Exome Sequencing Project cohort; ISR=McGill Israel cohort; FC=McGill French-Canadian cohort; NY=McGill Columbia University Spot Study; FE=fixed effects; $I^2$=I-square test for heterogeneity.

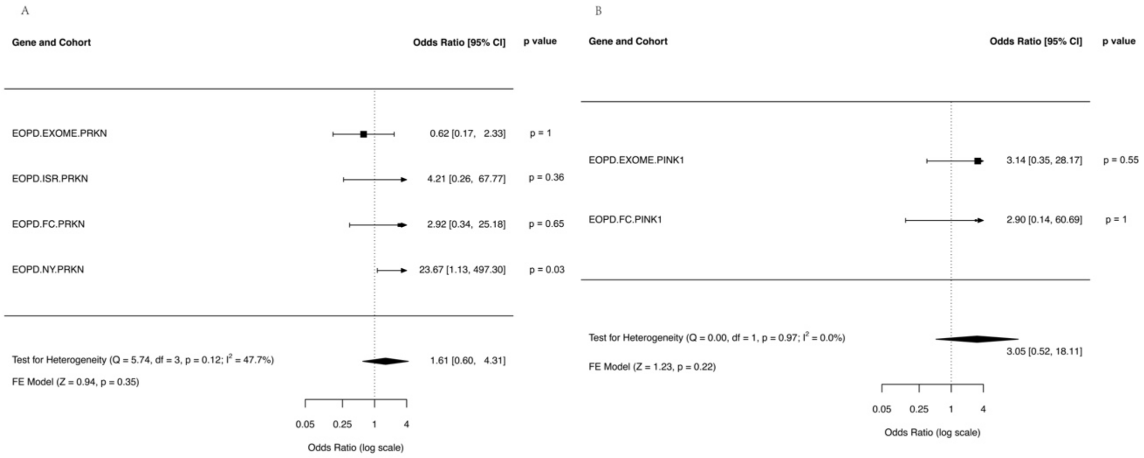


**Supplementary figure 3. Meta-analysis of carriership burden of private damaging variants in *PRKN and PINK1* genes in the EOPD cohort using allele count data**.

Forest plot of (A) *PRKN* (*Parkin*), (B) *PINK1* per cohort in rare private damaging variants group (CADD >20) in the EOPD cohort. Allele counts for each gene were extracted from the IPDGC Whole Exome Sequencing Project cohort and McGill cohort separately. Meta-analyses were performed using *Metafor*. Panels show Odds ratio, confidence intervals (CI), corrected Fisher's exact test gene burden p-value per cohort (right side of each plot), and the overall meta-analysis p-value per gene (bottom left of each plot).

Key: EOPD=Early-onset Parkinson's disease; EXOME=IPDGC Whole Exome Sequencing Project cohort; ISR=McGill Israel cohort; FC=McGill French-Canadian cohort; NY=McGill Columbia University Spot Study; FE=fixed effects; I²=I-square test for heterogeneity.

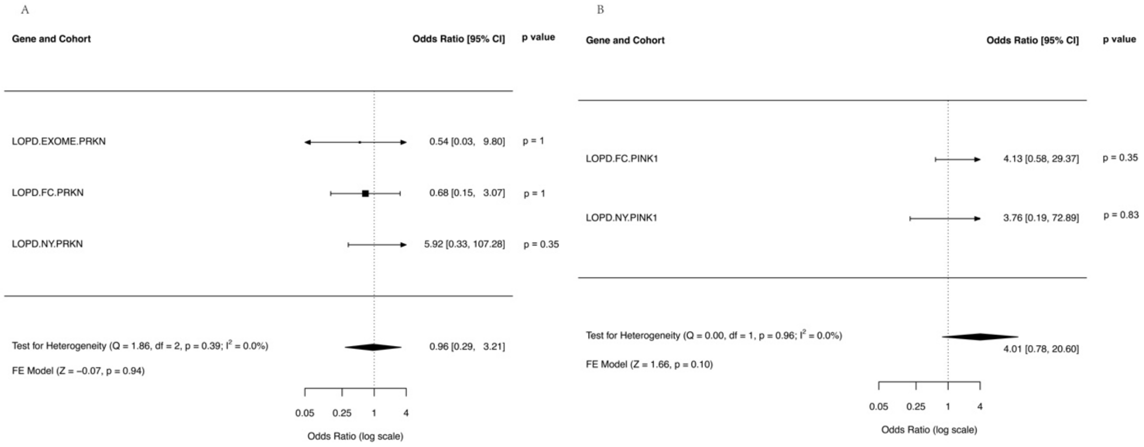


**Supplementary figure 4. Meta-analysis of carriership burden of private variants in *PRKN and PINK1* genes in the LOPD cohort using allele count data**.

Forest plot of (A) *PRKN* (*Parkin*), (B) *PINK1* per cohort in rare private variants group in the LOPD cohort. Allele counts for each gene were extracted from the IPDGC Whole Exome Sequencing Project cohort and McGill cohort separately. Meta-analyses were performed using *Metafor*. Panels show Odds ratio, confidence intervals (CI), corrected Fisher's exact test gene burden p-value per cohort (right side of each plot), and the overall meta-analysis p-value per gene (bottom left of each plot).

Key: LOPD=Late-onset Parkinson's disease; EXOME=IPDGC Whole Exome Sequencing Project cohort; FC=McGill French-Canadian cohort; NY=McGill Columbia University Spot Study; FE=fixed effects; $I^2$=I-square test for heterogeneity.

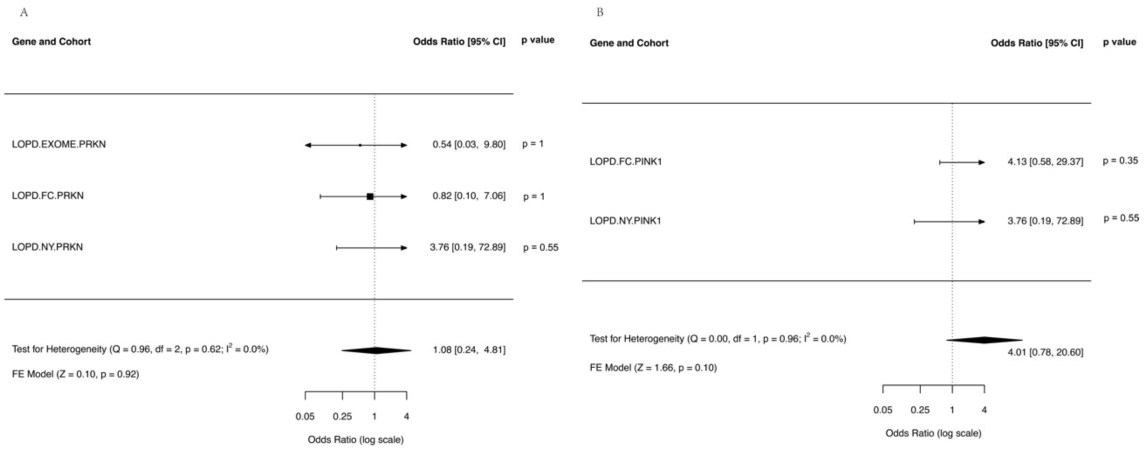


**Supplementary figure 5. Meta-analysis of carriership burden of private damaging variants in *PRKN and PINK1* genes in the LOPD cohort using allele count data**.

Forest plot of (A) *PRKN* (*Parkin*), (B) *PINK1* per cohort in rare private damaging variants group (CADD >20) in the LOPD cohort. Allele counts for each gene were extracted from the IPDGC Whole Exome Sequencing Project cohort and McGill cohort separately. Meta-analyses were performed using *Metafor*. Panels show Odds ratio, confidence intervals (CI), corrected Fisher's exact test gene burden p-value per cohort (right side of each plot), and the overall meta-analysis p-value per gene (bottom left of each plot).

Key: LOPD=Late-onset Parkinson's disease; EXOME=IPDGC Whole Exome Sequencing Project cohort; FC=McGill French-Canadian cohort; NY=McGill Columbia University Spot Study; FE=fixed effects; $I^2$=I-square test for heterogeneity.

**Supplementary Table 1. Overview of the cohort information for different tests**

| Method | Dataset | Data Type | PD | EOPD | LOPD | Controls | Total |
|---|---|---|---|---|---|---|---|
| SKAT-O Test | AMP-PD-Genome | WGS | 2,494 | 206 | 2,288 | 3,559 | 6,053 |
| | IPDGC-WES | WES | 1,108 | 584 | 76 | 456 | 1,564 |
| | McGill NY | MIPs | 893 | 92 | 801 | 428 | 1,321 |
| | McGill FC | MIPs | 696 | 154 | 542 | 2,275 | 2,971 |
| | McGill ISR | MIPs | 638 | 125 | 513 | 503 | 1,141 |
| Fisher's exact Burden test | PD Genome Project | WGS | 2,744 | - | - | 4,071 | 6,815 |
| | IPDGC Resequencing project | Resequencing data | 3,071 | - | - | 2,136 | 5,207 |
| | UK Biobank | WES | 114 | - | - | 38,263 | 38,377 |
| | IPDGC-WES | WES | 1,108 | 584 | 76 | 456 | 1,564 |
| | McGill NY | MIPs | 893 | 92 | 801 | 428 | 1,321 |
| | McGill FC | MIPs | 910 | 160 | 566 | 2,330 | 3,240 |
| | McGill ISR | MIPs | 661 | 125 | 513 | 523 | 1,184 |

WGS=Whole genome sequencing; WES=Whole exome sequencing; MIPs=Targeted next-generation sequencing with Molecular inversion probes; PD=Parkinson's disease; EOPD=Early-onset Parkinson's disease; LOPD=Late-onset Parkinson's disease; NY=McGill Columbia University Spot Study; FC=McGill French/French-Canadian cohort; ISR=McGill Israel cohort.

**Supplementary Table 2. Assessment of private variants in *PRKN, PARK7* and *PINK1* with Parkinson's disease (PD) in EOPD cohorts with available sequencing data.**

| Gene | Variant Group | *SKAT-O* (p-values) | | | | | *MetaSKAT* |
|---|---|---|---|---|---|---|---|
| | | AMP-PD | IPDGC-WES | ISR | FC | NY | Meta P-values (corrected) |
| *PRKN* | Private | 0.52 | 0.51 | 0.69 | 0.42 | 0.54 | 0.40 (1.00) |
| | Private damaging | 0.73 | 0.20 | 0.69 | 0.19 | 0.54 | 0.50 (1.00) |
| *PINK1* | Private | 0.13 | 0.33 | - | 0.60 | - | 0.65 (1.00) |
| | Private damaging | 0.30 | 0.33 | - | 0.60 | - | 0.72 (1.00) |
| *PARK7* | Private | 0.57 | 0.33 | - | - | - | 0.41 (1.00) |
| | Private damaging | 0.56 | - | - | - | - | 0.86 (1.00) |

Sequence-kernel-association test (*SKAT-O*) was used to assess variant burden for individual cohort analysis and *MetaSKAT* was used to perform the meta-analysis. Uncorrected P-values for the SKAT-O tests are indicated. Corrected P-values for the meta-analyses are indicated in parentheses.

Key: EOPD=Early-onset Parkinson's disease; AMP-PD=Accelerating Medicine Partnership in Parkinson's disease cohort; IPDGC-WES=International Parkinson's disease Genomics Consortium whole exome sequencing cohort; ISR=McGill Israel cohort; FC=McGill

French/French-Canadian cohort; NY=McGill Columbia University Spot Study.

**Supplementary Table 3. Assessment of private variants in *PRKN, PARK7* and *PINK1* with Parkinson's disease (PD) in LOPD cohorts with available sequencing data.**

| Gene | Variant Group | *SKAT-O* (p-values) | | | | | *MetaSKAT* |
|---|---|---|---|---|---|---|---|
| | | AMP-PD | IPDGC-WES | ISR | FC | NY | Meta P-values (corrected) |
| *PRKN* | Private | 0.55 | 0.39 | - | 0.88 | 0.24 | 0.90 (1.00) |
| | Private damaging | 0.57 | 0.39 | - | 0.94 | 0.36 | 0.89 (1.00) |
| *PINK1* | Private | 0.22 | - | - | 0.14 | 0.38 | 0.13 (0.39) |
| | Private damaging | 0.24 | - | - | 0.14 | 0.38 | 0.13 (0.39) |
| *PARK7* | Private | 0.57 | - | - | - | 0.73 | 0.74 (1.00) |
| | Private damaging | 0.53 | - | - | - | - | 0.50 (1.00) |

Sequence-kernel-association test (*SKAT-O*) was used to assess variant burden for individual cohort analysis and *MetaSKAT* was used to perform the meta-analysis. Uncorrected P-values for the SKAT-O tests are indicated. Corrected P-values for the meta-analyses are indicated in parentheses.

Key: LOPD=Late-onset Parkinson's disease; AMP-PD=Accelerating Medicine Partnership in Parkinson's disease cohort; IPDGC-WES=International Parkinson's disease Genomics Consortium whole exome sequencing cohort; ISR=McGill Israel cohort; FC=McGill

French-Canadian cohort; NY=McGill Columbia University Spot Study.

**Acknowledgments**

We would like to thank all of the subjects who donated their time and biological samples to be part of this study. We would also like to thank all members of the International Parkinson's Disease Genomics Consortium (IPDGC). For a complete overview of members, acknowledgments and funding, please see http://pdgenetics.org/partners. We would like to thank the Accelerating Medicines Partnership initiative (AMP-PD) for the publicly available whole-genome sequencing data. The AMP-PD cohort includes Michael J. Fox Foundation (MJFF) and National Institutes of Neurological Disorders and Stroke (NINDS) BioFIND study, Harvard Biomarkers Study (HBS), the NINDS Parkinson's disease Biomarkers Program (PDBP), MJFF Parkinson's Progression Marker Initiative (PPMI), NIA International Lewy Body Dementia Genetics Consortium Genome Sequencing in Lewy body dementia case-control cohort (LBD), the MJFF *LRRK2* Cohort Consortium (LCC) cohort, and the NINDS Study of Isradipine as a Disease Modifying Agent in Subjects With Early Parkinson Disease, Phase 3 (STEADY-PD3). A full list of PDBP investigators can be found at https://pdbp.ninds.nih.gov/policy. Data used in the preparation of this article were obtained from the PPMI database (www.ppmi-info.org/data). For up-to-date information on the study, visit www.ppmi-info.org. PPMI – a public-private partnership – is funded by the Michael J. Fox Foundation for Parkinson's Research funding partners 4D Pharma, Abbvie, Acurex Therapeutics, Allergan, Amathus Therapeutics, ASAP, Avid Radiopharmaceuticals, Bial Biotech, Biogen, BioLegend, Bristol-Myers Squibb, Calico, Celgene, Dacapo Brain Science, Denali, The Edmond J. Safra Foundation, GE Healthcare, Genentech, GlaxoSmithKline, Golub Capital, Handl Therapeutics, Insitro, Janssen Neuroscience, Lilly, Lundbeck, Merck, Meso Scale Discovery, Neurocrine Biosciences, Pfizer, Piramal, Prevail, Roche, Sanofi Genzyme, Servier, Takeda, Teva, UCB, Verily, and Voyager Therapeutics. Industry partners are contributing to PPMI through financial and in-kind donations and are playing a lead role in providing feedback on study parameters through the Partner Scientific Advisory Board (PSAB). Through close interaction with the study, the PSAB is positioned to inform the selection and review of potential progression markers that could be used in clinical testing. As registered users of the AMP-PD, JH, BIB, and SJL have access to individual-level data. We would like to thank the PD Variant Browser team who developed and contributed to this dataset and made it publicly available. The access to part of the participants for this research has been made possible thanks to the Quebec Parkinson's Network (http://rpq-qpn.ca/en/).

**Conflicts of Interests**

Z.G.O is on the Scientific Advisory Board of Bial Biotech Inc. and Handl Therapeutics, and received consultancy fees from Bial Biotech Inc., Handl Therapeutics (UCB), Denali, Neuron23, Ono Therapeutics, Prevail Therapeutics, Idorsia, Guidepoint and Lighthouse. D.K. is the Founder and Scientific Advisory Board Chair of Lysosomal Therapeutics Inc. and Vanqua Bio. D.K. serves on the scientific advisory boards of The Silverstein Foundation, Intellia Therapeutics, AcureX and Prevail Therapeutics and is a Venture Partner at OrbiMed. J.H., C.H.W., D.S., E.A.F., E.Y., F.A., L.K., P.S., R.N.A., S.H.B., B.R.Z., B.I.B. and S.J.L. declare that they have no competing interests.

**Funding**

The analysis of cohorts at McGill University was financially supported by grants from the Michael J. Fox Foundation, the Canadian Consortium on Neurodegeneration in Aging (CCNA), the Canada First Research Excellence Fund (CFREF), awarded to McGill University for the Healthy Brains for Healthy Lives initiative (HBHL), and Parkinson Canada. Z.G.O. is supported by the Fonds de recherche du Québec - Santé (FRQS) Chercheurs-boursiers award and is a William Dawson Scholar. D.K. is supported by the Simpson Querrey Center for Neurogenetics. The research of C.H.W. is supported by Neuraly, the consulting fee is supported by Kyowa, Sunovion, Acadia and Abbvie, and the speaker's honoraria is supported by Adamas, Amneal, Kyowa and Neurocrine.

**Code availability**

https://github.com/ipdgc/IPDGC-Trainees/blob/master/Private-variants%20data%20processing.md

**Data availability**

All genetic data used in this study is available (upon application) from the following sites:

(i) Accelerating Medicines Partnership: Parkinson’s Disease (https://amp-pd.org).

(ii) International Parkinson’s Disease Genomics Consortium (https://pdgenetics.org/resources).

(iii) Parkinson's Disease Variant Browser (https://pdgenetics.shinyapps.io/VariantBrowser).

(iv) Dataset using targeted next-generation sequencing with Molecular inversion probes (https://cbigr-open.loris.ca/)
